# Severe visceral leishmaniasis in Ethiopia: outcomes, co-infections and mortality in a prospective real-world cohort

**DOI:** 10.64898/2025.12.19.25342642

**Authors:** Eleni Ayele, Saskia van Henten, Desalew Getahun Ayalew, Saba Atnafu, Asnakew Engidaw Mereed, Hana Yohannes, Tigist Mekonnen, Tadfe Bogale, Aman Mossa, Gebrehiwot Lema, Jemal Yasin, Nicole Berens-Riha, Thao-Thy Pham, Carl Boodmann, Annelies Mondelaers, Wim Adriaensen, Saïd Abdellati, Ermias Diro, Rezika Mohammed, Fabiana Alves, Mezgebu Silamsaw Asres, Myrthe Pareyn, Mekibib Kassa, Johan van Griensven

**Author notes:** Corresponding author Eleni Ayele, MD Department of Internal Medicine, University of Gondar Gondar, Ethiopia.

## Abstract

**Background:** Visceral leishmaniasis (VL) remains a major public health challenge in East Africa, particularly in Ethiopia. Clinical trials on VL often exclude patients with severe comorbidities or laboratory abnormalities, limiting the generalizability of evidence used to guide real-world management. We comprehensively characterised the clinical profile, treatment outcomes, and mortality of VL patients treated in a referral centre.

**Methododology/Principal findings:** This prospective cohort study enrolled patients at the Leishmaniasis Research and Treatment Center (LRTC), University of Gondar (02/2023-06/2024). All VL cases fulfilling eligibility criteria underwent detailed clinical, laboratory, and radiological assessments. VL treatment followed WHO guidelines. Outcomes, adverse events, and supportive care measures were recorded during treatment and over a 12-month follow-up period. Patients meeting at least one exclusion criterion (e.g. comorbidities, clinical signs of severe VL) of standard phase III VL treatment trials were defined as trial-ineligible patients and compared with trial-eligible patients. A total of 314 VL patients, mostly young adult males (median age 22 years), were enrolled. Of these, 21 (6.7%) had HIV co-infection; 204 (65%) met ≥1 key exclusion criterion typically used in VL clinical trials. Trial-ineligible patients had high parasitemia, more concurrent infections, more pronounced clinical and laboratory abnormalities and a higher need of supportive care measures including systemic antibiotics and blood transfusion. Overall cure rate was 90.1%. Mortality after the first VL treatment course was 6.4%, ranging from 1.8% in trial-eligible patients to 8.8% in trial-ineligible patients. Leading causes of death included severe bacterial infections, acute liver failure, and haemorrhagic complications.

**Conclusions:** These findings underscore the severity and complexity of VL in routine care in our study population, and highlight the limitations of current trial populations to inform broader clinical practice. Strengthening supportive care and expanding research inclusivity designed to respond to the needs of this patient population are critical to achieve VL elimination targets.

**Author Summary:** Visceral leishmaniasis is a life-threatening disease that continues to affect many people in East Africa, especially in Ethiopia. However, many of the patients we see in everyday clinical practice are very different from the carefully selected patients who take part in clinical trials. Trials often exclude people who are severely ill or who have other medical problems, which means that the results may not fully represent the patients who most need treatment. In this study, we followed people treated for visceral leishmaniasis at a referral centre to better understand how they present, how they respond to treatment, and what challenges they face. Most of the patients we treated would not have been eligible for typical clinical trials. These patients were more severely ill, had more infections, and required more intensive supportive care, such as antibiotics or blood transfusions. While most people were cured, those with more severe illness had a higher risk of death, often due to serious infections or bleeding. By describing the real-world situation of patients with visceral leishmaniasis, we show that current research does not fully capture the needs of the most vulnerable groups. We highlight the importance of improving supportive care and making future studies more inclusive.

## Introduction

Visceral leishmaniasis (VL) is a disseminated protozoan infection transmitted by sandflies. It is caused by intracellular parasites belonging to the *Leishmania (L.) donovani* complex. Globally, around 50,000-90,000 cases are estimated to occur annually [1]. VL is caused by zoonotic *L. infantum* in Latin America and the Mediterranean region, and by anthroponotic *L. donovani* in the Indian subcontinent and East-Africa. Currently, East-Africa carries 73% of the global VL burden, with Sudan, South Sudan and Ethiopia most affected [2].

The World Health Organization (WHO) recently launched a framework for the elimination of VL as a public health problem, with reducing case fatality in primary VL to below 1% as a main indicator in the Neglected Tropical Diseases (NTD) road map [3]. Ethiopia, one of the highest burden countries for VL in East Africa, plays a critical role in these elimination efforts [4,5]. The elimination strategy emphasizes early diagnosis, effective treatment, and comprehensive patient management to reduce transmission and mortality as one of the main pillars to achieve VL elimination.

However, reducing case fatality requires prompt access to diagnosis and treatment, but also a clear understanding of the clinical and laboratory features of VL patients in routine care. This knowledge is crucial for improving patient care and outcomes, thereby contributing to reduced mortality—an essential component of the elimination strategy. In East-Africa, studies describing clinical and laboratory features in detail are mostly clinical trials, which usually involve highly selected patient populations and controlled environments, while data on more general patient populations are often limited to basic socio-demographic variables. As such, available information may not fully reflect the realities of standard clinical care. Hence, data from routine hospital care, where patients present with a wide range of comorbidities and varying degrees of disease severity, are essential for identifying key shortcomings in care provision and guide the design of interventions adapted for the different patient populations [5,6].

This prospective cohort study aims to fill this critical gap by comprehensively describing clinical and laboratory features of VL in a tertiary hospital in North-West Ethiopia, and characterizing the prevalence of common co-infections and co-morbidities in VL-endemic regions. Additionally, we provide an overview of the supportive care provided and describe the main causes of death. We also compare the profile, management and outcomes of typical clinical trial patients versus those generally excluded from clinical trials.

## Methods

### Study setting

The study was conducted at the Leishmania Research and Treatment center (LRTC) at the University of Gondar, North-West Ethiopia. The center, supported by the University of Gondar, the Drugs for Neglected Diseases Initiative (DND*i*), and the Institute of Tropical Medicine (ITM) conducts clinical trials on VL and provides free health care to routine VL patients. LRTC serves as the key referral center for treatment of VL in the Amhara region since 2004 [6]. Most VL cases treated at the center are seasonal migrant workers, traveling from various parts of the country to the VL-endemic lowland cash crop farming areas of North-West Ethiopia. Additionally, the center treats residents of these lowland areas and patients referred from other centers for further diagnostic work-up or due to severe disease [7].

### Study design and population

We conducted a prospective cohort study including all VL cases treated at LRTC between February 2023 and June 2024. We excluded patients below the age of 12 years, those already on VL treatment for three days or more, and those unable to give blood or come for follow-up. Over the planned recruitment period of 18 months, we expected to recruit 741 patients from LRTC and the Abdurafi health center supported by Médecins Sans Frontières (MSF), but due to insecurities in the area the study was only conducted at LRTC. The study was registered on ClinicalTrials.gov (identifier NCT05602610).

### Routine VL diagnosis and treatment

Individuals presenting at the University of Gondar Comprehensive Specialized Hospital and meeting the VL clinical case definition (fever for two or more weeks and enlarged spleen or either loss of weight, anemia or leukopenia) with a travel history to or residents of a VL endemic area were considered as VL suspects and referred to LRTC for diagnostic assessment. In each primary VL case (no previous VL), rK39 rapid diagnostic test (RDT) (IT-LEISH, Bio-Rad, USA) and spleen aspiration was done. In case of contraindications to spleen aspiration (*e.g.* platelets<40,000 or signs of increased bleeding tendency), bone marrow aspiration was performed. For those with previous VL history (*i.e.* cases of relapse), only tissue aspiration was done. A VL diagnosis was made if *Leishmania* parasites were detected microscopically in the tissue aspirate. If no parasites were detected, a diagnosis of primary VL could (occasionally) still be made based on the combination of high clinical suspicion and a positive rK39 RDT result. All VL cases were also tested for HIV co-infection.

VL treatment was provided according to the national guideline [8]. Most primary VL was treated with a combination of sodium stibogluconate (SSG) (20 mg/kg intramuscular (IM) or intravenous (IV) with a maximum dose of 850 mg) and paromomycin (PM) (11 mg/kg IM) for 17 days. Liposomal amphotericin B (L-AmB; AmBisome) (30mg/kg IV over 12 days) was reserved for complicated VL cases, including those with VL relapse, or older age (> 45 years) and more severe VL symptoms (*e.g.* critical condition, severe laboratory abnormalities). In case of HIV co-infection, L-AmB 30 mg/kg was combined with miltefosine (MF; 2.5 mg/kg per os (PO) daily for 28 days) for 1 or 2 cycles. All patients were admitted for monitoring, with regular clinical and laboratory assessments during the treatment period. Treatment response was assessed at the end of treatment visit (day 17 for SSG/PM and AmB- and day 28 for L-AmB/MF-treated patients) and was based on clinical signs (such as absence of fever, improved appetite, weight gain, and reduced spleen size) and laboratory results (improvement in hematologic profile). If there was no significant clinical improvement and no change in hematologic profile, or in case of HIV coinfection, a parasitological test of cure was performed by microscopic evaluation of spleen or bone marrow tissue aspirates for the detection of parasites using Giemsa staining. If parasites remained detectable, the patient was defined to have treatment failure and additional treatment was given. If there was suspicion of relapse during the follow-up period after treatment, VL was confirmed by spleen or bone marrow tissue aspiration except for HIV coinfected patients with chronic, relapsing VL who receive treatment based on clinical symptoms, as they did not manage to clear parasites with treatment anymore.

### Investigations, data recording and patient follow-up

After VL diagnosis, a comprehensive clinical evaluation was conducted and data were collected on VL-related symptoms/signs and clinical parameters. Blood, stool and urine was collected for further analysis, and electrocardiogram (ECG) (GE Healthcare, Mac 2000, India) and chest X-ray (DogX-ray, China) were performed. If there was a productive cough, sputum was collected and sent for tuberculosis (TB) testing using GeneXpert (Cepheid, USA). In case of clinical suspicion of disseminated TB, an abdominal ultrasound (Samsung Medison, Republic of Korea) was performed.

During treatment, clinical and laboratory assessments were performed on day 3, day 7, and at the end of treatment study visit. After discharge, follow-up visits were planned for three and six months after the end of treatment for clinical and laboratory evaluation. Phone calls were made on month 9 and month 12 after admission to document the long-term outcomes. Individuals missing planned study visits were also contacted by phone and invited for a clinical assessment at LRTC or, if not able to attend, to collect information on their VL-related health. In case of a relapse and subsequent treatment, a post-relapse visit after 6 months was scheduled. The study closed at the end of June 2025.

We documented the anti-*Leishmania* treatment given, the occurrence of adverse events, reasons for treatment change and the response to treatment. Ancillary and supportive care were documented as well, including all concomitant treatments received with indications. The need of advanced supportive care (e.g. intensive care, dialysis, Fluid and ionotropic use…) and transfusion of blood products (e.g. whole blood, fresh frozen plasma, platelets) was also recorded. Patients were given antibiotics on indication, based on the suspected focus. Prescriptions of antibiotics was not informed by CRP-levels, as this was a study-related procedure. For patients with severe neutropenia (an absolute neutrophil count <500 cells/ µL), antibiotics were given (ciprofloxacin PO or ceftriaxone IV) for prophylaxis or presumed sepsis treatment. In case an individual died, the main cause of death was defined by the treating clinician. To avoid directing the treating clinician towards certain terms or causes of death, the information was entered using free text without pre-defined categories for causes of death or contributing factors.

### Laboratory assays and radiological investigations

The following laboratory tests were performed on blood at baseline: a complete blood count (SysmexXN350 and SX500, Japan), kidney and liver function tests, liver enzymes (Humastar 200, Germany), electrolytes (Humalyte Plus Human Germany), C-Reactive Protein (CRP) (Quick Read Go instrument, Finland), International Normalized Ratio (INR; Coagucheck XS, Germany) and HIV serology (One Step Anti-HIV (1 & 2), China). For *Leishmania*, besides the routine rK39 RDT and microscopy of Giemsa-stained spleen or bone marrow aspirates, real-time PCR targeting kinetoplast DNA (kDNA) on whole blood was done in batches as described before [9].

Additional tests included urine dipstick and microscopy, Giemsa-stained thick and thin blood films for malaria and stool wet mount microscopy for intestinal parasite eggs and trophozoites (Lugol’s iodine and normal saline wet mount). For chest X-ray and abdominal ultrasound analyses, we used the routine report from the radiologist. Based on the abdominal ultrasound report, the treating VL clinician – extensively experienced in VL care - defined whether the radiological findings were compatible with disseminated TB.

### Data collection and statistical analysis

The study was conducted in compliance with Good Clinical Practice (GCP). Data were collected electronically using the GCP compliant REDCap software Demographic, clinical and laboratory features of the patients were summarized in descriptive analysis. Continuous variables were reported as medians (interquartile range), while binary/categorical variables were expressed as frequencies and percentages. Where relevant, continuous variables were categorized using clinically relevant cut-offs. Analysis was done using Stata version 18 (StataCorp LP, College Station, TX, USA).

Treatment outcomes were described after the first course of treatment (at the end of treatment visit) and at discharge from hospital (end of the VL episode), also taking into consideration additional treatment for treatment failures. Outcomes at the end of treatment and at discharge were defined as cure, treatment failure, deceased, defaulted or referred. The number and proportion of patients with relapse after discharge was reported as well.

To define the group of patients that would typically be excluded from clinical trials (“trial-ineligible patients”), we applied the revised trial exclusion criteria from the phase III clinical trial comparing MF/PM and SSG/PM, conducted by DND*i* at seven sites in Kenya, Uganda, Sudan, and Ethiopia, including at the LRTC in Gondar [10]. In this trial, the original (stricter) exclusion criteria were revised during the study to allow for recruitment of a more representative VL population. In the present paper, we will refer to this trial as the MF/PM trial [10]. The revised exclusion criteria we applied were age <4YRS or >45yrs, relapse, HIV coinfection, concurrent TB, clinically significant ECG abnormalities, haemoglobin <5 g/dL and severe VL according to the investigator’s judgement, requiring AmBisome therapy (see Table S1).

We summarized the information provided by the treating clinician on the main cause of death for each individual, grouped by the main underlying process or organ affected (*e.g.* severe infection or acute liver failure). The cause(s) of death for each individual can be found in Annex Table S2. Only deaths occurring during hospitalization for VL treatment were recorded. Hence individuals known to have died after discharge outside the hospital were not included in Table S2.

### Ethics

The study protocol was approved by the institutional review board of the Institute of Tropical Medicine Antwerp, Antwerp, Belgium (ref 1544/21) and the University of Gondar, Ethiopia (ref VP/RTT/05/965/2022), the ethical committee of the University of Antwerp, Belgium (ref 3276) and the Ethiopian national research ethics review committee (ref 17/152/147/23). Written informed consent was obtained from all participants.

## Results

### Baseline characteristics, medical history and diagnostic work-up

During the recruitment period, a total of 916 VL suspects were seen, of which 558 were diagnosed with VL (Fig. 1). Of these, 454 VL cases were treated at LRTC, of which 314 (69.2%) were included in this study. A total of 140 VL cases were not included, due to inclusion in another study, unable to adhere to the follow-up visits, unable or refusing to provide informed consent and having started *Leishmania* treatment for more than 3 days.

**Figure 1.**
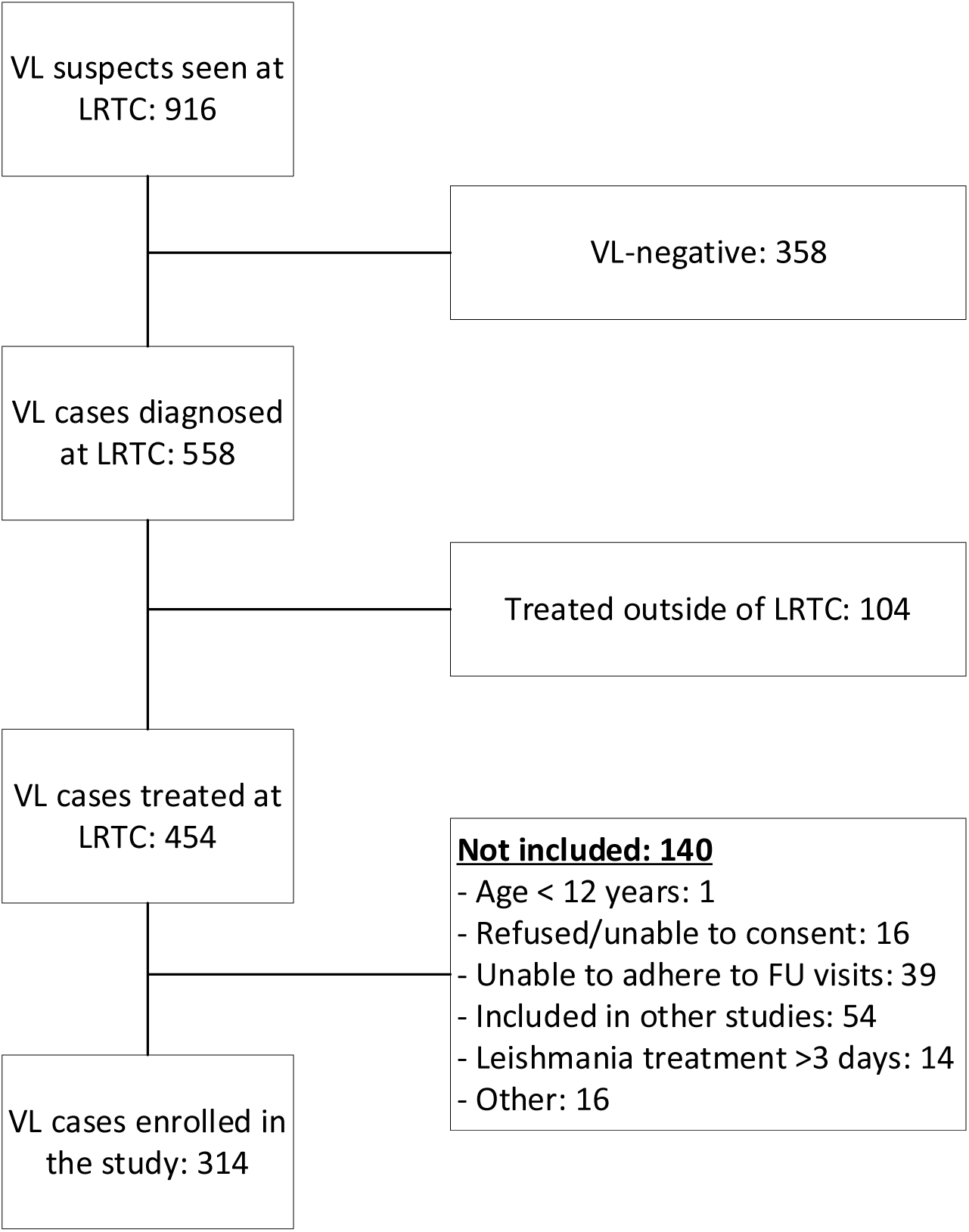
Flowchart describing the recruitment in the visceral leishmaniasis (VL) cohort study in Ethiopia, 2023-2024. LRTC: Leishmaniasis Research and Treatment Center. FU: follow-up.

All but one of the 314 study participants included were male; the median age was 22 years (IQR 20-28) (Table 1). The median duration of symptom onset was 41 days (IQR 30-70); only 71 (22.7%) participants presented within 30 days of symptom onset. Twenty (6.4%) participants were previously diagnosed with HIV, of which all were on antiretroviral treatment (ART) and four were on TB treatment. Fourteen (4.5%) participants had previously been treated for VL. Almost half (47%) of the participants reported to have taken traditional medicine for the current disease episode prior to attending LRTC.

**Table 1.** Baseline characteristics, medical history, VL diagnosis and treatment at the LRTC, Gondar, Ethiopia (2023-2024)

| Variable; median (IQR) or n (%) | All<br>(n=314) | Trial-eligible<br>(n=110) | Trial-ineligible<br>(n=204) |
| --- | --- | --- | --- |
| Age, years | 22 (20-28) | 21 (19-25) | 21 (19-25) |
| Male sex | 313 (99.6) | 110 (100) | 203 (99.7) |
| Rural residence | 279 (88.8) | 102 (92.7) | 177 (86.8) |
| Travel history to endemic area or<br>migrant worker | 273 (86.9) | 99 (90.0) | 174 (85.3) |
| Traditional medicine used before VL | 147 (47) | 46 (41.8) | 101 (49.8) |
| On TB treatment at enrolment | 4 (1.3) | 0 (0) | 4 (2.0) |
| **HIV history** |  |  |  |
| Previously diagnosed with HIV | 20 (6.4) | 0 (0) | 20 (9.8) |
| On ART at enrolment | 20 (6.4) | - | 20 (9.8) |
| Time on ART, years | 4.8 (0.6-10.6) | - | 4.8 (0.6-10.6) |
| CD4 count, cells/μL (n=13) | 98 (52-114) | - | 98 (52-114) |
| **VL history** |  |  |  |
| Previous VL episode | 14 (4.5) | - | 14 (6.9) |
| Number of previous VL episodes | 4.5 (2-9) | - | 4.5 (2-9) |
| Months since last VL episode | 10 (4-12) | - | 10 (4-12) |
| **VL diagnosis<sup>a</sup>** |  |  |  |
| Tissue aspirate positive | 293 (93.3) | 110 (100) | 183 (89.7) |
| rK39 RDT+/ tissue aspirate negative | 14 (4.5) | 0 (0) | 14 (6.9) |
| rK39 RDT+/ tissue aspirate not done | 2 (0.6) | 0 (0) | 2 (1.0) |
| rK39 RDT-/tissue aspirate negative | 1 (0.3) | 0 (0) | 1 (0.5) |
| Chronic, relapsing VL in HIV patients<sup>b</sup> | 4 (1.3) | 0 (0) | 4 (2.0) |
| **VL treatment<sup>c</sup>** |  |  |  |
| Sodium stibogluconate/paromomycin | 217 (69.3) | 110 (100) | 107 (52.7) |
| AmBisome | 73 (23.3) | 0 (0) | 73 (36.0) |
| AmBisome & miltefosine | 20 (6.4) | 0 (0) | 20 (9.8) |
| Miltefosine/paromomycin | 3 (1.0) | 0 (0) | 3 (1.5) |

VL: visceral leishmaniasis; ART: antiretroviral treatment; RDT: rapid diagnostic test; AmBisome: liposomal amphotericin B; TB: tuberculosis; IQR: interquartile range
<sup>a</sup> Detailed diagnostic procedure provided in Annex Results S1.
<sup>b</sup> In patients with multiple relapses and whereby parasitological cure can generally not be achieved anymore, tissue aspiration is not performed for diagnosis of VL relapse but treatment is started on clinical grounds.
<sup>c</sup> One patient with severe VL died before VL treatment could be started

With the revised MF/PM trial exclusion criteria, 204 (65.0%) of the 314 cases met at least one exclusion criterion and were considered “trial-ineligible patients” while 110 (35%) were “trial-eligible patients”.

Symptoms and clinical signs,

Fever (97.4%), weight loss (95.5%), edema (48.7%), vomiting (22.0%), diarrhea (22.3%), dyspnea (24.9%), cough (69.4%) and an increased bleeding tendency (24.5%) were commonly reported by the patients (Table 2). On clinical examination at baseline, oedema (36.9%), ascites (14.0%), jaundice (15.9%), anemia (75.5%), spontaneous bleeding (6.4%) were commonly observed. 39% had severe weakness or were in a state of collapse. On clinical examination, abnormal respiratory findings (mainly crepitation and absent air entry) were found in 14%, and abnormal cardiac findings in 8.0%, including a gallop rhythm in 11 (3.5%). The median body mass index was 16.9 kg/m^2^. Several findings were more common in ‘trial-ineligible patients’, such as the presence of cough, diarrhea, ascites, jaundice, oedema, bleeding tendencies, respiratory or cardiac abnormalities on examination, and severe weakness at enrolment (Table 2).

**Table 2.** Symptoms and clinical findings in VL cases at LRTC, Gondar, Ethiopia (2023-2024)

| Symptoms, n (%) | All (n=314) | Trial-eligible<br>(n=110) | Trial-ineligible<br>(n=204) |
| --- | --- | --- | --- |
| Fever | 305 (97.4) | 109 (100) | 196 (96.1) |
| Weight loss | 297 (95.5) | 107 (98.2) | 190 (94.1) |
| Loss of appetite | 260 (82.8) | 89 (80.9) | 171 (83.8) |
| Dyspnea | 78 (24.9) | 23 (21.1) | 55 (27.0) |
| Cough | 218 (69.4) | 71 (64.6) | 147 (72.1) |
| Vomiting | 69 (22.0) | 22 (20) | 47 (23.0) |
| Diarrhea | 70 (22.3) | 19 (17.3) | 51 (25.0) |
| Abdominal pain | 137 (43.6) | 41 (37.3) | 96 (47.1) |
| Oedema | 153 (48.7) | 23 (20.9) | 130 (63.7) |
| Spontaneous bleeding | 77 (24.5) | 9 (8.2) | 68 (33.3) |
| **Clinical findings, n (%)** |  |  |  |
| Ascites | 44 (14.0) | 3 (2.7) | 41 (20.1) |
| Jaundice | 50 (15.9) | 0 (0) | 50 (24.5) |
| Oedema | 116 (36.9) | 0 (0) | 116 (56.9) |
| Anemia | 237 (75.5) | 73 (66.4) | 164 (80.4) |
| Lymphadenopathy | 9 (2.9) | 0 (0) | 9 (4.4) |
| Spontaneous bleeding sites |  |  |  |
| Skin | 8 (2.6) | 0 (0) | 8 (3.9) |
| Mucosa | 12 (6.0%) | 0 (0) | 12 (5.8) |
| General condition |  |  |  |
| No or limited weakness | 191 (61.0) | 87 (79.1) | 104 (51.2) |
| Severe weakness<sup>a</sup> | 116 (36.9) | 23 (20.9) | 93 (45.6) |
| State of collapse<sup>b</sup> | 6 (1.9) | 0 (0) | 6 (2.9) |
| Liver size (cm) | 11 (10-12) | 11 (10-12) | 11 (10-12) |
| Spleen size (cm) | 8 (5-10) | 7 (5-10) | 8 (5-10) |
| Body mass index (kg/m<sup>2</sup>) | 16.9 (15.8-17.8) | 17.0 (15.8-17.6) | 16.7 (15.8-18.0) |
| Cardiac findings on examination | 25 (8.0) | 2 (1.8) | 23 (11.3) |
| Gallop rhythm | 11 (3.5) | 0 (0) | 11 (5.4) |
| Systolic murmur | 14 (4.5) | 2 (1.8) | 12 (5.9) |
| Abnormal respiratory clinical findings | 44 (14.0) | 5 (4.6) | 39 (19.0) |

VL: visceral leishmaniasis <sup>a</sup>difficulty walking unsupported; <sup>b</sup>unable to stand from sitting position

### Laboratory and radiological evaluations

Laboratory results are shown in Table 3. While hematological abnormalities were common, severe abnormalities were also seen in around a third of patients such as hemoglobin < 7 g/dL in 25.2%, platelets < 20.000/µL in 9.3% and severe neutropenia in 37.6%. Severe abnormalities in liver enzyme tests such as alanine transaminase five times above the limit of normal occurred in 11.8% of the cases, while creatinine levels were generally normal. Hypokalemia was seen in 48% cases and hyponatremia in 92.0%. Albumin levels were decreased in all but one patient. INR was increased in all but 11 patients, and above 1.5 in 76 (24.3%); eight individuals had INR values > 8. CRP-levels were increased (≥ 3 mg/L) in all but 4, with a median of 42 mg/L (IQR 19-79), ranging from 1.9 up to 302. Levels > 100 mg/L were seen in 56 (18.1%) and < 10 mg/L in 40 (12.9%). *Leishmania* blood PCR was positive in 299 (95.2%), with a median cycle threshold (Ct) value of 23 cycles (IQR 21-27).

**Table 3.**
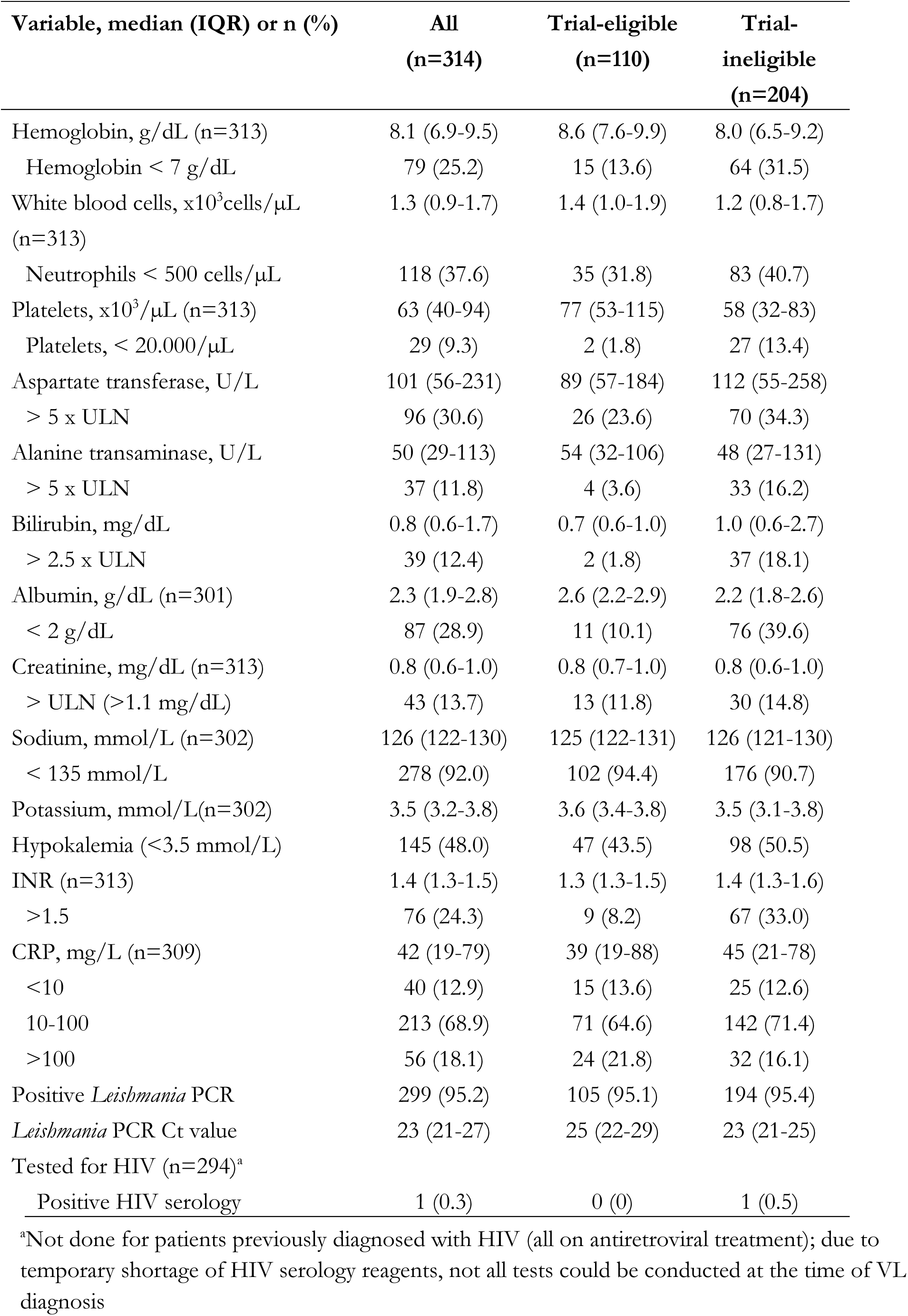
Baseline blood analysis results of VL cases in Gondar, Ethiopia (2023-2024)

### Co-infections

Amongst the 294 individuals tested for HIV infection at the time of VL diagnosis, one was newly diagnosed with HIV during the study period (Table 3). Combined with the 20 previously diagnosed HIV patients (all on ART), the prevalence of VL-HIV coinfection was 6.7% (21/314). Two patients had *Plasmodium falciparum* parasites identified microscopically in their blood. Stool microscopy detected parasites in 117 (38.6%) participants. Using urine microscopy, hematuria was found in 13%, proteinuria in 53.5% and pyuria in 4.4% (Table 4).

**Table 4.**
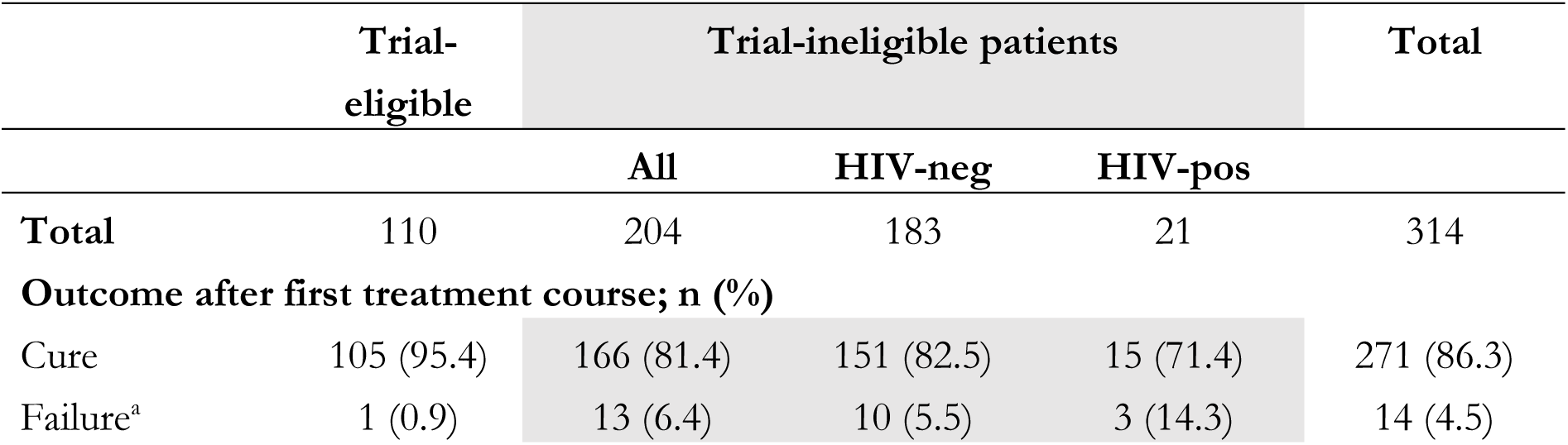
Microscopy, radiological and other investigations for concurrent conditions or complications in VL cases in Gondar, Ethiopia (2023-2024)

Chest X-ray analyses showed that pneumonia was diagnosed in 49 (17.8%) participants, and TB was reported to be likely in one. Other abnormalities included pleural effusions and cardiomegaly.

Abdominal ultrasound was done based on clinical suspicion of TB in 130 patients. Based on the ultrasound findings, disseminated TB was considered likely in 23 (17.7%) participants.

Most laboratory and radiological abnormalities were more common in trial-ineligible patients, including severe thrombocytopenia, high INR values (including all eight with values > 8), severe liver test abnormalities, severe hypoalbuminemia and chest X-ray abnormalities.

### VL treatment and treatment outcomes

Overall, 217 (69.3%) patients with VL were started on SSG/PM and 93 (29.7%) on L-AmB-based treatment, the latter exclusively in trial-ineligible patients (Table 1). Three were started on MF/PM, as L-AmB was not available. One patient with severe VL died before VL treatment could be initiated. After the first VL treatment course, 271 (86.3%) were cured and 14 (4.5%) individuals had treatment failure, of which 12 were cured after additional treatment, one was referred to another center and one defaulted. At discharge (outcomes of the VL episode, including additional therapy for initial failures), 283 (90.1) were cured, 24 (7.6%) had died, six (1.9%) had defaulted and one (0.3%) was referred (Table 5).

**Table 5.**
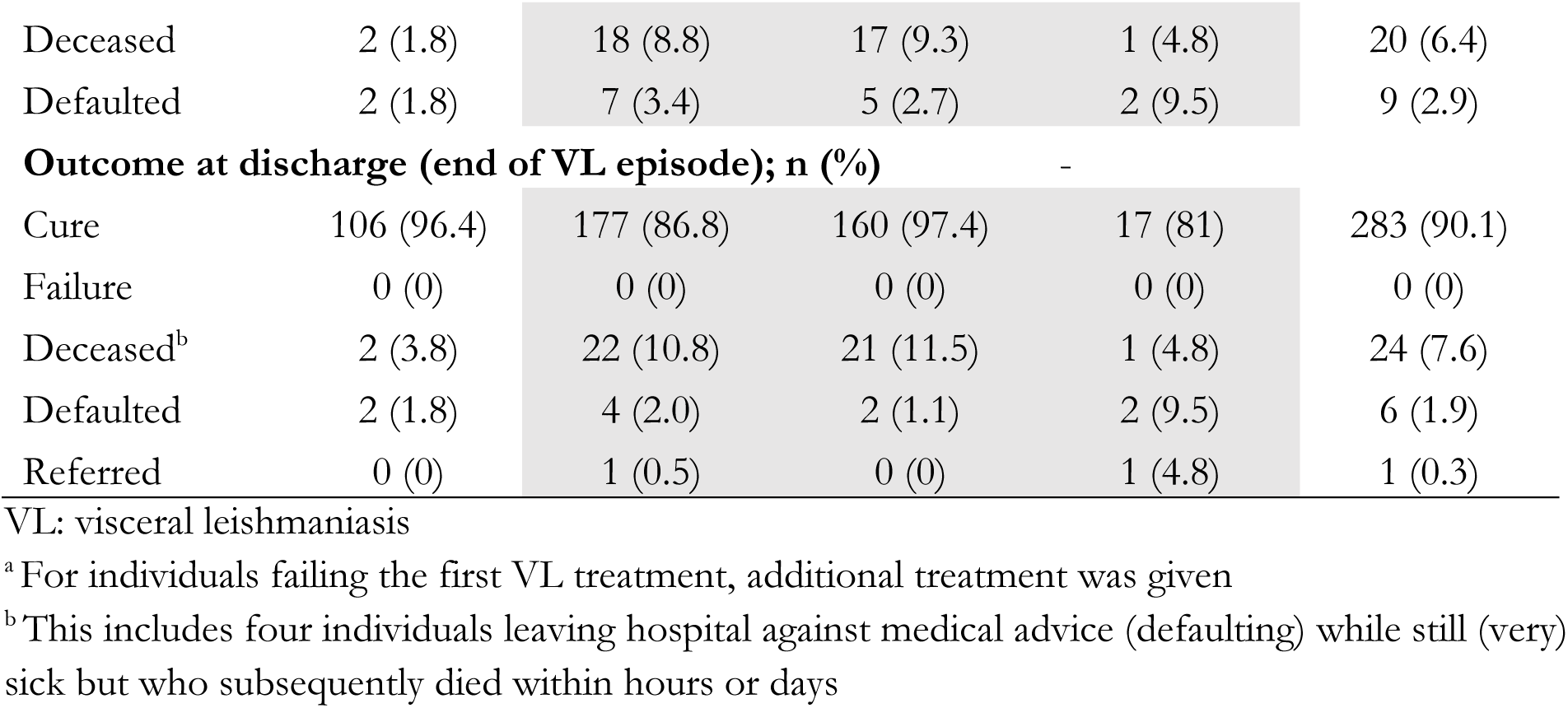
Treatment outcomes of VL cases in Gondar, Ethiopia (2023-2024)

Mortality (10.8%) and failure rates (6.4%) were higher in the trial-ineligible patients. While mortality was highest (11.5%; 21/183) in HIV-negative trial-ineligible patients, failure rates were relatively high in HIV patients (14.3%; 3/21). In primary VL cases, mortality was 7.7% (23/300).

Treatment was changed due to treatment-related adverse events in 17 (5.4%) participants, of which 16 individuals had received SSG/PM. Amongst these, the main adverse events leading to treatment change were due to renal (n=8), liver (n=6) and cardiac toxicity (n=2). One individual started on MF/PM was changed to L-AmB due to worsening of liver enzymes. None of the 93 patients started on L-AmB-based therapy had to change treatment due to drug-related adverse events.

Outcome information was available by six months for 191 (60.8%) and by 12 months for 133 (43.4%) participants. After the VL episode, 14 relapses were documented during follow-up, of which two died during treatment. Six were seen in non-HIV-coinfected individuals and ten relapses occurred within six months after treatment. Three patients died during follow-up due to events apparently not related to VL (one due to acute watery diarrhea, one due to meningitis, one due to a gun shot). Overall, 29 (9.2%) of the 314 study participants were reported to have died.

### Use of antibiotics and other supportive care measures

Out of the 314 study participants, systemic therapeutic antibiotics were given to 156 (49.7%) during the treatment period. 113 (36.0%) individuals received therapeutic antibiotics within 48 hours of VL diagnosis and 81 (25.8%) later during admission (Table 6). The most common indication was pneumonia, with otitis media, meningitis, urinary, gastro-intestinal, and skin and soft tissue infections contributing to a smaller extent. While ceftriaxone (with or without azithromycin) was most commonly used (36.3%), ceftazidime/vancomycin combination therapy was not uncommon (10.5%).

**Table 6.** Therapeutic use of systemic antibiotics, type and indications in VL cases in Gondar, Ethiopia (2023-2024)

| Variable, n (%) | All<br>(n=314) | Trial-eligible<br>(n=110) | Trial-ineligible<br>(n=204) |
| --- | --- | --- | --- |
| Therapeutic antibiotics started | 156 (49.7) | 38 (34.5) | 118 (57.8) |
| Within 48 hours of VL diagnosis | 113 (36.0) | 26 (26.4) | 84 (41.2) |
| After 48 hours of VL diagnosis | 81 (25.8) | 14 (12.7) | 67 (32.8) |
| Main indications |  |  |  |
| Pneumonia | 134 (42.7) | 30 (27.3) | 104 (51.0) |
| Urinary/gastro-intestinal infection | 19 (5.9) | 7 (6.4) | 12 (5.9) |
| SSTI/otitis media | 8 (2.5) | 2 (1.8) | 6 (2.9) |
| Meningitis | 2 (0.6) | 0 (0) | 2 (1.0) |
| Most common antibiotics prescribed |  |  |  |
| Ceftriaxone & azitromycin | 71 (22.6) | 15 (13.6) | 56 (27.4) |
| Ceftriaxone | 43 (13.7) | 13 (11.8) | 30 (14.7) |
| Ceftazidime & vancomycin | 33 (10.5) | 5 (4.5) | 28 (13.7) |
| Ciprofloxacin | 22 (5.1) | 6 (5.4) | 16 (7.8) |
| Amoxicillin/clavulanic acid +/-<br>azitromycin | 5 (1.6) | 2 (1.8) | 3 (1.5) |
| Prophylactic antibiotics<sup>a</sup> | 62 (19.7) | 23 (20.9) | 39 (19.1) |

<sup>a</sup> Prophylactic antibiotics (ciprofloxacin po or ceftriaxone iv) were given to patients with an absolute neutrophil count <500 cells/ μL
VL: visceral leishmaniasis; SSTI: skin and soft tissue infections

Antibiotic prescription was frequent across all baseline CRP categories, with little apparent discrimination by CRP level. Among participants with CRP <10 mg/L at baseline, 32.5% received antibiotics within 48 hours of VL diagnosis, while 37.5% of those with CRP >100 mg/L received antibiotics in the same time window (Table S3). As CRP testing was done solely for study purposes and was not available to treating clinicians, these data suggest that early antibiotic initiation was largely independent of the degree of systemic inflammation as reflected by CRP.

TB was diagnosed during the VL episode in 15 (4.8%) patients, which was defined as disseminated TB in 11, pulmonary TB in two, central nervous system TB in one, and pericardial TB in one. Only in two cases, the diagnosis was bacteriologically confirmed by sputum analysis.

Twenty (6.4%) study participants were transfused with whole blood transfusion, ten (3.2%) received platelets and 13 (4.1%) fresh frozen plasma (Table 7). All but one blood products were administered to trial-ineligible patients. Potassium supplementation was given to 125 (39.8%) predominantly to trial-ineligible patients and vitamin K to 20 (6.4%), exclusively to trial-ineligible patients. Inotropes were prescribed to 9 (2.9%); corticosteroids were given to 19 (6.1%). Ten (3.2%) were referred to the intensive care unit but for five transfer was not possible (due to financial or logistical reasons).

**Table 7.** Main treatment and care beside VL treatment and antibiotics provided to VL cases in Gondar, Ethiopia (2023-2024)

| Anti-infectious<br>treatment, n (%) | All<br>(n=314) | Trial-eligible<br>(n=110) | Trial-ineligible<br>(n=204) |
| --- | --- | --- | --- |
| TB treatment<sup>a</sup> | 19 (5.1) | 0 (0) | 19 (7.8) |
| Antimalarials | 24 (7.6) | 7 (6.4) | 17 (8.3) |
| **Other treatment** |  |  |  |
| Costicosteroids | 19 (6.1) | 1 (0.9) | 18 (8.8) |
| Inotropics/vasopressor | 9 (2.9) | 0 (0) | 9 (4.4) |
| Vitamin K | 20 (6.4) | 0 (0) | 20 (9.8) |
| K supplementation | 125 (39.8) | 24 (21.8) | 101 (49.5) |
| **Blood products** |  |  |  |
| Whole blood | 20 (6.4) | 0 (0) | 20 (9.8) |
| Platelets | 10 (3.2) | 1 (0.9) | 9 (4.4) |
| Fresh frozen plasma | 13 (4.1) | 0 (0) | 13 (6.4) |
| **Intensive care (IC)** |  |  |  |
| Indication for IC | 10 (3.2) | 1 (0.9) | 9 (4.4) |
| Dialysis indicated | 3 | 0 | 3 |
| Dialysis done<sup>b</sup> | 0 | 0 | 0 |

<sup>a</sup> Four patients were taking TB treatment at time of VL diagnosis, 15 started during VL treatment
<sup>b</sup> Not possible as patients could not afford

### Causes of death

Table 8 summarizes the information on the main cause of death for 24 patients that died while residing in hospital for VL treatment, as provided on the death report by the treating physician (see Table S2 of Annex for the information listed for each individual). Severe bacterial infections - with or without other conditions - were mentioned in 13 patients, predominantly pneumonia. Acute liver failure was defined as cause of death in eight individuals. Acute renal failure was mentioned for four cases, and severe bleeding disturbances for three. Two patients died presumably due to fatal pulmonary embolism.

**Table 8.**
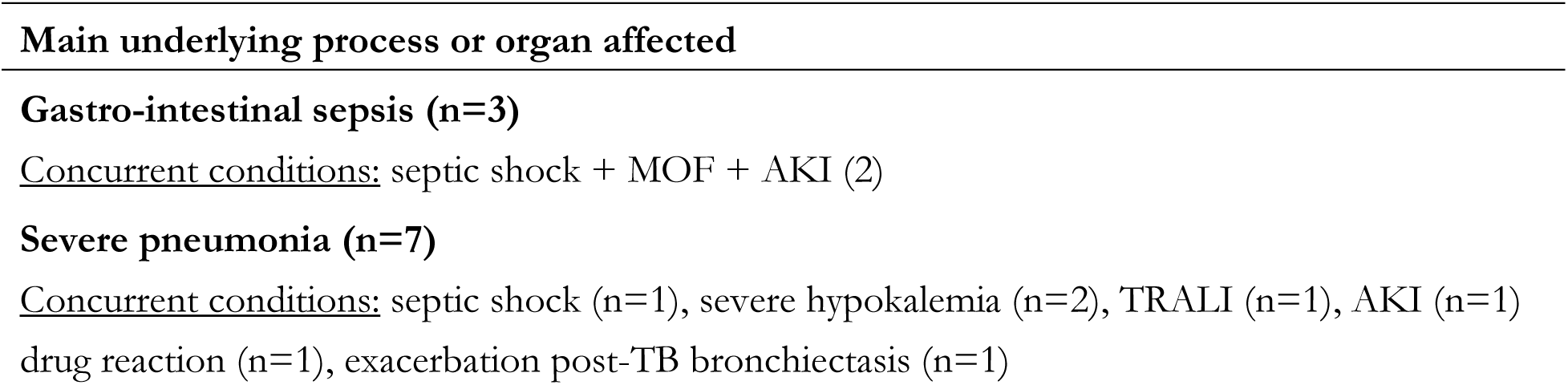
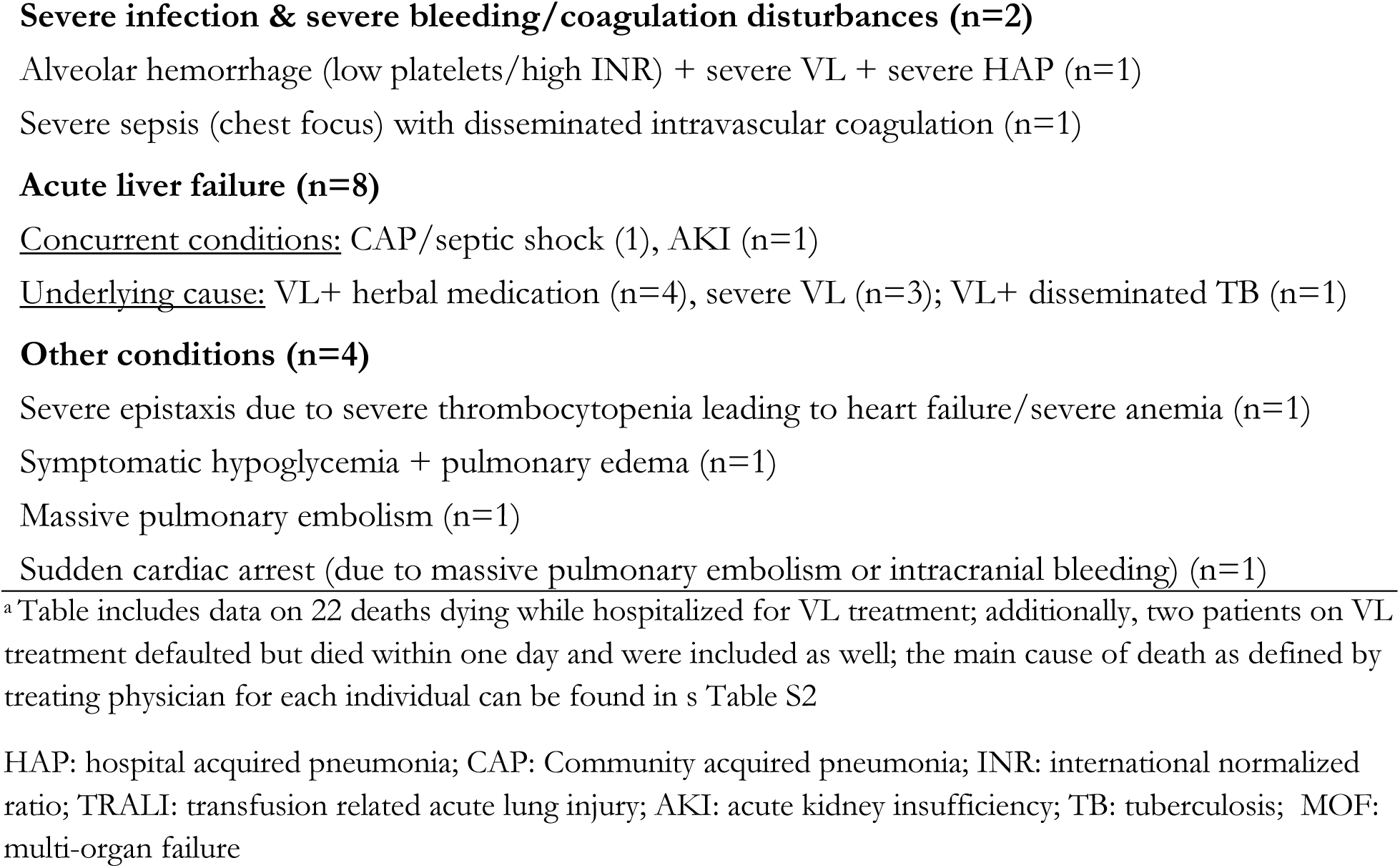
Main causes of death in VL cases in Gondar, Ethiopia (2023-2024)^a^

## Discussion

In this VL cohort, including VL cases across the full spectrum of severity, a wide range of organs and systems were affected, some leading to pronounced pulmonary, hepatic, renal and coagulation dysfunction and abnormalities. The need for supportive care was high, including high therapeutic use of antibiotics, cardiovascular support and transfusion of blood products. Important reported causes of death included severe bacterial infection (particularly pulmonary), liver and renal failure, fatal hemorrhages and cardiac arrest. As the most comprehensive characterization of VL in East-Africa, this study can provide avenues for improving supportive care in VL and define areas for future research.

Patients with varying degrees of VL severity were included in this study. Importantly, the majority (65%) of VL cases would typically not be included in phase III clinical trials, especially those with signs of severe disease - requiring AmBisome therapy - and HIV co-infected patients. Mortality was highest in HIV-negative trial-ineligible patients, at 11.5%. Phase III clinical trials are designed to identify safe and effective antileishmanial treatments, but not to inform the evidence-base for improved management of complicated VL cases. With the ambition to reduce case fatality to below 1% and to decentralize VL care, this gap will have to be filled. Additional research is needed to optimize VL care for severe VL cases, to define which patients can be safely managed at the lower health care level, which patients would require referral to secondary and tertiary care levels, and what package of care needs to be available at the different levels of health care facilities.

Despite antileishmanial treatment, VL can potentially progress to multi-organ failure resembling sepsis. Some authors have suggested that VL could drive a protracted sepsis-like syndrome - referred to as leishmanial sepsis - mediated by dysregulated immune response which further increases the susceptibility to invasive bacterial infections and bacterial sepsis further aggravating the inflammatory process [11]. In the same study site, we previously found culture-proven bloodstream infections in 19% of VL cases [11,12].

In our cohort, severe bacterial infections were the leading cause of death, being reported in 54% (13/24) of deceased participants. This is consistent with data from Brazil, where bacterial infection was also the main cause of death in VL, followed by bleeding, liver failure, and renal failure [13].

Together, these observations highlight that timely recognition and treatment of bacterial co-infection is critical to prevent clinical deterioration and death in VL.

Appropriate antibiotic use in VL is, however, challenging because clinical manifestations of VL and bacterial sepsis overlap. In most patients in our study, therapeutic antibiotics were initially withheld, as the presenting fever was often judged to be VL-related. In this context, host inflammatory markers such as CRP could be valuable adjuncts to guide decisions on when to start antibiotics, particularly when the clinical picture is equivocal.

CRP and other acute phase proteins are likely upregulated in VL itself [13,15–17], and the optimal thresholds to indicate concomitant bacterial infection remain uncertain [14]. Nevertheless, baseline CRP values in our cohort showed considerable variability, and antibiotic prescribing did not meaningfully track this variation. With clinicians unaware of the CRP results, 32.5% of patients with CRP <10 mg/L received early antibiotics, whereas 62.5% of those with CRP >100 mg/L did not.

These patterns suggest that, if prospectively validated thresholds were available, CRP (alone or combined with other host biomarkers) might help reduce unnecessary antibiotic use in patients with very low CRP values and, conversely, prompt earlier targeted treatment among patients with markedly elevated CRP.

Further studies are therefore needed to define CRP- and biomarker-based algorithms for antibiotic initiation in VL and to evaluate their impact on patient outcomes and antimicrobial stewardship.

Such tools may be particularly useful in lower-level health facilities, where VL expertise and diagnostic capacity are limited. In better resourced centers, systematic use of blood cultures could complement biomarker-based approaches by identifying the causative pathogen and its antibiotic susceptibility.

Spontaneous bleeding was commonly observed on admission, and some patients died due to fatal bleedings. To what extent the increased bleeding tendency in VL cases is related to low or dysfunctional platelets, coagulation disturbances or both remains poorly understood [13]. In our study, INR values were increased in 96.5% of patients, but rarely markedly. Baseline thrombocytopenia was common, but < 20.000/µL occurred in less than 10%. Outside of Africa, data suggest that disseminated intravascular coagulation occurs in most patients [18]. While often subclinical, overt clinical manifestations with pronounced bleeding have been reported as well [19–21]. Liver dysfunction, vitamin K malabsorption and sepsis could potentially further worsen coagulation disturbances [13]. In our center, vitamin K supplements, fresh frozen plasma and/or platelets are routinely given (if available) in cases of severe laboratory abnormalities or overt bleeding. Although the evidence-base supporting such practices remains limited, particularly with ongoing disseminated intravascular coagulation, it might be prudent to refer severe VL cases with spontaneous bleeding to referral centers capable of administering blood products.

In line with other studies, liver disturbances were common, often pronounced and liver failure was considered the main cause of death in eight individuals [22,23]. While hepatic effects of VL have been consistently reported [13,21,24], other factors could further exacerbate this, including use of traditional medicine, sepsis and concurrent medication given during admission [13]. This warrants careful use of potentially hepatotoxic medication, with close laboratory monitoring, access to AmBisome for those with pronounced liver dysfunction, and early identification of sepsis. A similar approach could apply to renal dysfunction. While rare on admission, it required treatment change in eight individuals and contributed to death in several cases. As herbal medicine can be liver or nephrotoxic [25], more work is needed to understand the types of herbal medicine being used in VL endemic areas and how it affects the course and management of VL.

In summary, strategies to prevent clinical deterioration and reduce case fatality should focus on careful evaluation for bacterial infections and early antibiotic use, effective measures for infection prevention and control, judicious use of potentially toxic concurrent medication with close monitoring, cardiovascular support, and careful administration of blood products. With two deaths potentially related to fatal pulmonary embolism, the role of prophylactic anticoagulation should be further explored, despite the increased bleeding tendency commonly seen in VL [26,27].

Strengths of the study are that it was conducted in a highly experienced *Leishmania* treatment and research center, with a comprehensive laboratory assessment supported by high-quality laboratory facilities. Additionally, we included the full spectrum of VL cases, in contrast with clinical trials, and hence more representative of the overall VL population seeking care in an East-African setting.

There are a number of important limitations to acknowledge. Not all patients diagnosed with VL at the center could be included. Fifty-three patients were enrolled in other studies of which 21 are included in a clinical trial, which could over-represent the “trial-ineligible” patients in the present study. It also needs to be acknowledged that LRTC is a referral center receiving complicated cases, and it may not fully represent other treatment centers managing VL. Some tests such as additional coagulation tests or blood culture could have been of value. Due to the ongoing conflict and instability in the region during the study, long-term outcomes were not available for a substantial proportion of study participants.

In conclusion, our study shows that at the main VL treatment center in Ethiopia, most VL patients present with severe disease, with multiple organs affected. Severe bacterial infections, severe bleeding and liver failure were commonly diagnosed and potentially leading to death. Most of our patients would typically not be included in clinical trials, and this group displayed high mortality.

Besides strategies aiming at early VL diagnosis and treatment, enhanced VL care will be needed to decrease case fatality rates, the main indicator of success for the East-African VL elimination initiative.

## Annex – Supporting information

Results S1. Additional information on the diagnosis of visceral leishmaniasis

Table S1. Overview of the main exclusion criteria from the miltefosine/paromomycin trial that were used to define the non-trial group within the study populationa

Table S2. Line list of the main causes of death in VL cases in Gondar, Ethiopia (2023-2024)

Table S3. Prescription of systemic therapeutic antibiotics within 48 hours of VL diagnosis, stratified by CRP values at baseline in VL cases in Gondar, Ethiopia (2023-2024)

## Author contributions

**Conceptualization:** Eleni Ayele, Rezika Mohammed, Johan Van Griensven, Saskia Van Henten

**Methodology:** Saskia Van Henten, Annelies Mondelaers, Said Abdellati, Wim Adriaensen, Ermias Diro

**Investigation:** Eleni Ayele, Desalew Getahun Ayalew, Gebrehiwot Lema Legese, Mezgebu Silamsaw Asres, Jemal Yasin, Aman Mossa, Hana Yohannes, Mekbib Kassa, Asnakew Engidaw Meered, Saba Atnafu, Tigist Mekonnen, Hana Yohannes, Thao-Thy Pham

**Data curation:** Tadfe Bogale, Hana Yohannes, Eleni Ayele, Saskia Van Henten, Johan Van Griensven, Nicole Berens-Riha

**Formal analysis:** Eleni Ayele, Nicole Berens-Riha, Johan Van Griensven

**Visualization:** Johan Van Griensven, Nicole Berens-Riha

**Supervision:** Eleni Ayele, Saskia Van Henten, Fabiana Alves, Johan Van Griensven

**Project administration:** Eleni Ayele, Tadfe Bogale, Saskia Van Henten, Johan Van Griensven

**Funding acquisition:** Myrthe Pareyn, Johan Van Griensven

**Writing – original draft:** Eleni Ayele, Johan Van Griensven **Writing – review & editing:** All authors

## Data Availability

All relevant data are in the manuscript and its supporting information files.

## Annex - Supplementary Information

### Results

#### Results S1. Additional information on the diagnosis of visceral leishmaniasis

Tissue aspiration was done for 308 at initial diagnosis. In four patients, it was not done because these were well-known HIV positive patients where after multiple previous VL episodes, parasite clearance could not be achieved anymore and it was decided to start treatment in case of clinical worsening. For two primary VL cases, there were contraindications for splenic aspiration and no bone marrow set was available, hence treatment was started based on a positive rK39 RDT result. All these six cases without tissue aspirate had a positive *Leishmania* blood PCR result at enrolment. Out of 308 undergoing tissue aspiration, 293 were positive. The 15 patients with a negative bone marrow (n=14) or spleen aspirate (n=1) were primary VL cases started on treatment based on a positive rK39 RDT (n=14) or clinical suspicion (n=1). Twelve of these cases were subsequently found to have a positive *Leishmania blood* PCR result.

---

**Table S1.** Overview of the main exclusion criteria from the miltefosine/paromomycin trial that were used to define the trial-ineligible group within the study population**^a^**

| **Revised (looser) exclusion criteria** |  |
| --- | --- |
|  | No parasitological confirmation of VL on tissue aspiration |
|  | PKDL grade 3 on VL diagnosis |
|  | Any anti-*Leishmania* drugs < 6 months |
|  | Age < 4 or > 50 years old |
|  | VL history (previous VL episode) |
|  | HIV co-infection |
|  | Concurrent tuberculosis |
|  | Female patients of child bearing age not accepting a pregnancy test and/or not agreeing to use contraception |
|  | Hemoglobin < 5 g/dL |
|  | Severe VL according to the treating physician, based on clinical manifestations (such as jaundice, bleeding, oedema)<sup>b</sup> and/or clinically significant laboratory abnormalities<sup>c</sup> |
|  | Clinically significant ECG abnormalities at baseline |
| VL: visceral leishmaniasis; AmB; PKDL: post-kala azar dermal leishmaniasis |  |

<sup>a</sup> For Ethiopia specifically, exclusion criteria for malnutrition in adults were based on skin-fold measurements, which were not available in CPS.
<sup>b</sup> All those with an indication for AmBisome defined by the treating physician based on clinical/laboratory severity indicators were considered trial-ineligible patients; additionally, those with jaundice, oedema, or epistaxis on clinical examination or reporting recent ( $\leq 7$ days) onset epistaxis on admission were considered trial-ineligible patients (even if AmBisome treatment would not be indicated), as was done during the miltefosine/paromomycin trial.
<sup>c</sup> The following tests were taken into account to define clinically significant laboratory abnormalities: haemoglobin, platelets, liver enzymes, total bilirubin, and creatinine

**Table S2.**
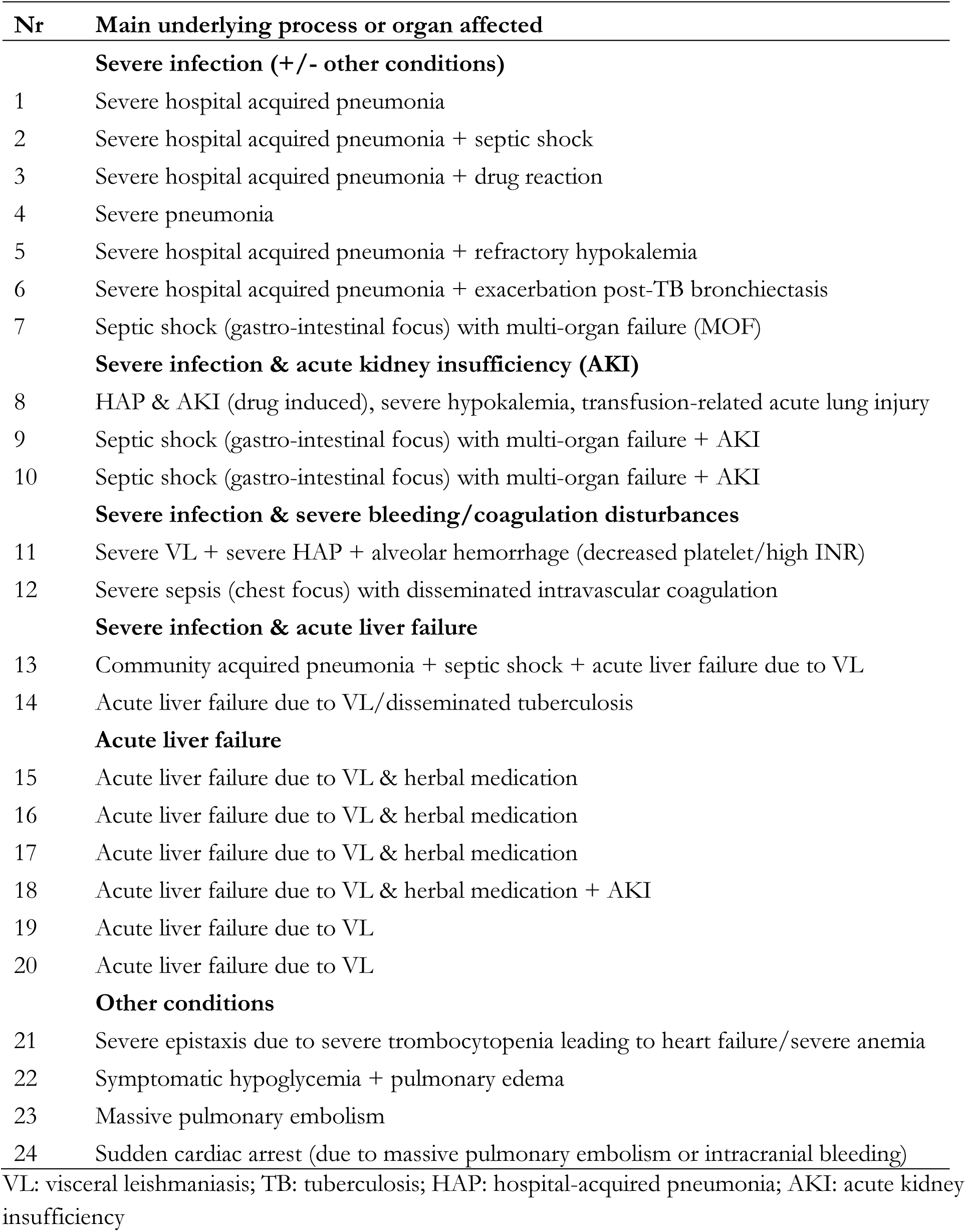
Line list of the main causes of death in VL cases in Gondar, Ethiopia (2023-2024)

**Table S3.** Prescription of systemic therapeutic antibiotics within 48 hours of VL diagnosis, stratified by CRP values at baseline in VL cases in Gondar, Ethiopia (2023-2024)

| CRP values at VL diagnosis<sup>a</sup> | Antibiotics prescribed (< 48 hours of VL diagnosis) |
| --- | --- |
| <10 mg/L | 13/40 (32.5%) |
| 10-49.9 mg/L | 41/127 (32.3%) |
| 50-99.9 mg/L | 35/86 (40.7%) |
| >100 mg/L | 21/56 (37.5%) |

<sup>a</sup> As CRP testing was done for study purposes, CRP values were not used by the treating physician when deciding on starting antibiotics
CRP: C-reactive protein; VL: visceral leishmaniasis

## References

1. World Health Organization. Global leishmaniasis surveillance, 2022: assessing trends over the past 10 years. Weekly Epidemiological Record. 2023;98(40):471–487. Available: https://www.who.int/publications/i/item/who-wer9840-471-487 Accessed 28 Oct 20252.

2. Federal Ministry of Health, Ethiopia. The Third National Neglected Tropical Diseases Strategic Plan 2021–2025. Addis Ababa: FMoH; 2021. Available: https://espen.afro.who.int/tools-resources/documents/third-national-neglected-tropical-diseases-strategic-plan-2021-2025 Accessed 28 Oct 2025.

3. World Health Organization. Strategic framework for the elimination of visceral leishmaniasis as a public health problem in Eastern Africa, 2023–2030. Geneva: World Health Organization; 2023. Available: https://www.who.int/publications/i/item/9789240094208. Accessed 28 Oct 2025

4. Deribe K, Meribo K, Gebre T, Hailu A, Ali A, Aseffa A, et al. The burden of neglected tropical diseases in Ethiopia, and opportunities for integrated control and elimination. Parasites & Vectors. 2012;5:240. doi:10.1186/1756-3305-5-240

5. World Health Organization. Global leishmaniasis surveillance update, 2024: consolidating gains and new initiatives. Weekly Epidemiological Record. 2025;100(45):535–552. Available: https://www.who.int/publications/i/item/who-wer10045-535-552. Accessed 28 Oct 2025.

6. Diro E, Lynen L, Mohammed R, Boelaert M, Hailu A, van Griensven J. High parasitological failure rate of visceral leishmaniasis to sodium stibogluconate among HIV co-infected adults in Ethiopia. PLoS Neglected Tropical Diseases. 2014;8(5):e2875. doi:10.1371/journal.pntd.0002875.

7. Coulborn RM, Gebrehiwot TG, Schneider M, Gerstl S, Adera C, Herrero M, et al. Barriers to access to visceral leishmaniasis diagnosis and care among seasonal mobile workers in Western Tigray, Northern Ethiopia: a qualitative study. PLoS Neglected Tropical Diseases. 2018;12(11):e0006778. doi:10.1371/journal.pntd.0006778.

8. Federal Ministry of Health (Ethiopia). Guideline for diagnosis, treatment and prevention of leishmaniasis in Ethiopia. Addis Ababa: FMoH; 2013. Available: . Accessed 28 Oct 2025.

9. van Griensven J, van Henten S, Kibret A, Kassa M, Beyene H, Abdellati S, et al. Prediction of visceral leishmaniasis development in a highly exposed HIV cohort in Ethiopia based on Leishmania infection markers: results from the PreLeisH study. EBioMedicine. 2024;110:105474. doi:10.1016/j.ebiom.2024.105474

10. Younis BM, Mudawi Musa A, Monnerat S, Abdelrahim Saeed M, Awad Gasim Khalil E, Elbashir Ahmed A, et al. Safety and efficacy of paromomycin/ miltefosine/liposomal amphotericin B combinations for the treatment of post-kala-azar dermal leishmaniasis in Sudan: A phase II, open label, randomized, parallel arm study. PLoS Neglected Tropical Diseases. 2023;17(8):e0011780. doi:10.1371/journal.pntd.0011780.

11. Endris M, Takele Y, Woldeyohannes D, Tiruneh M, Mohammed R, Moges F, et al. Bacterial sepsis in patients with visceral leishmaniasis in Northwest Ethiopia. BioMed Research International. 2014;2014:361058. doi:10.1155/2014/361058

12. Endris M, Takele Y, Woldeyohannes D, Unakal C, Moges F, Tiruneh M, et al. Characteristics of bacterial sepsis among patients with visceral leishmaniasis. Asian Pacific Journal of Tropical Biomedicine. 2014;4(11):871–875. doi:10.12980/APJTB.4.2014C1133.

13. Costa CHN, Chang KP, Costa DL, Cunha FVM. From infection to death: an overview of the pathogenesis of visceral leishmaniasis. Pathogens. 2023;12(7):969. doi:10.3390/pathogens12070969.

14. van Griensven J, Cnops L, de Weggheleire A, Declercq S, Bottieau E. Point-of-care biomarkers to guide antibiotic prescription for acute febrile illness in Sub-Saharan Africa: Promises and caveats. Open Forum Infectious Diseases. 2020;7(8):ofaa260. doi:10.1093/ofid/ofaa260.

15. Wasunna KM, Raynes JG, Were JBO, Muigai R, Sherwood J, Gachihi G, Carpenter L, McAdam KPWJ. Acute phase protein concentrations predict parasite clearance rate during therapy for visceral leishmaniasis. Transactions of the Royal Society of Tropical Medicine and Hygiene. 1995;89(6):678–681. doi:10.1016/0035-9203(95)90442-5.

16. Kip AE, Balasegaram M, Beijnen JH, Schellens JHM, de Vries PJ, Dorlo TPC. Systematic review of biomarkers to monitor therapeutic response in leishmaniasis. Antimicrobial Agents and Chemotherapy. 2015;59(1):1–14. doi:10.1128/AAC.04298-14.

17. Cerón JJ, Pardo-Marín L, Caldin M, Furlanello T, Solano-Gallego L, Tecles F, Bernal L, Baneth G, Martínez-Subiela S. Use of acute phase proteins for the clinical assessment and management of canine leishmaniosis: general recommendations. BMC Veterinary Research. 2018;14(1):196. doi:10.1186/s12917-018-1524-y.

18. Costa DL, Rocha RL, Carvalho RMA, Lima-Neto AS, Harhay MO, Costa CHN, et al. Serum cytokines associated with severity and complications of kala-azar. Pathogens and Global Health. 2013;107(2):78–87. doi:10.1179/2047773213Y.0000000078.

19. Huang DB, Wu JJ, Hamill RJ. Reactive hemophagocytosis associated with the initiation of highly active antiretroviral therapy (HAART) in a patient with AIDS. Scandinavian Journal of Infectious Diseases. 2004;36(7):516–519. doi:10.1080/00365540410020569

20. de Santana Ferreira E, de Souza Júnior VR, de Oliveira JFS, Costa MFH, da Conceição de Barros Correia M, de Sá AF. Rare association of consumptive coagulopathy in visceral leishmaniasis: A case report. Trop Doctor. 2021;51: 120–122. doi:10.1177/0049475520967239

21. El Hag IA, Hashim FA, El Toum IA, Homeida M, El Kalifa M, El Hassan AM. Liver morphology and function in visceral leishmaniasis (Kala-azar). Journal of Clinical Pathology. 1994;47(6):547–551. doi:10.1136/jcp.47.6.547.

22. Hailu W, Weldegebreal T, Hurissa Z, Tafes H, Omollo R, Yifru S, et al. Safety and effectiveness of meglumine antimoniate in the treatment of Ethiopian visceral leishmaniasis patients with and without HIV co-infection. Transactions of the Royal Society of Tropical Medicine and Hygiene. 2010;104(11):706–712. doi:10.1016/j.trstmh.2010.07.007.

23. Kager PA, Rees PH, Manguyu FM, Bhatt KM, Hockmeyer WT, Wellde BT, Lyerly WH Jr. Clinical presentation of visceral leishmaniasis in Kenya: a prospective study of 64 patients. Tropical and Geographical Medicine. 1983;35(4):323–331. PMID: 6670114.

24. Duarte MIS, Corbett CEP. Histopathological patterns of the liver involvement in visceral leishmaniasis. Revista do Instituto de Medicina Tropical de São Paulo. 1987;29(3):131–136. doi:10.1590/S0036-46651987000300003.

25. Teshager NW, Amare AT, Tamirat KS, Zeleke ME, Taddese AA. Traditional herbal medicine use doubled the risk of multi-organ dysfunction syndrome in children: A prospective cohort study. PLoS ONE. 2024;19(6):e0286233. doi:10.1371/journal.pone.0286233.

26. Gürkan E, Ünsal C, Başlamışlı F, Arslan D. Budd–Chiari syndrome associated with visceral leishmaniasis and factor V Leiden mutation. Journal of Thrombosis and Thrombolysis. 2004;18(3):205–207. doi:10.1007/s11239-005-0347-4

27. Balikci C, Ural K. Evaluation of cardiopulmonary biomarkers during different stages of canine visceral leishmaniasis. Revista MVZ Córdoba. 2018;23(2):6403–6413. doi:10.21897/rmvz.1236

